# A self-guided digital health application for LDL-cholesterol reduction in adults with hypercholesterolaemia (The DIGICHOL randomised controlled trial)

**DOI:** 10.64898/2026.09.15.26363103

**Authors:** Antje Riepenhausen, Clara Ekerdt, Björn Meyer, Kamila Jauch-Chara

## Abstract

Structured lifestyle modification is the guideline-mandated foundation of lipid management but is rarely delivered in routine care. We evaluated *lipodia*, a fully self-guided digital health application providing structured lifestyle support, in a pragmatic randomised controlled trial conducted online. Adults with LDL-cholesterol above their risk-adapted target (n = 278) were randomised 1:1 to usual care plus lipodia or usual care alone. After six months, lipodia significantly reduced LDL-cholesterol relative to usual care (adjusted between-group difference in relative change −3.9%; p = 0.029), with nearly twice as many users achieving a clinically meaningful reduction of at least 10% (41.7% versus 23.8%; p = 0.002). lipodia also significantly improved patient activation, non-HDL-cholesterol, and health-related quality of life, with high uptake, low attrition, and no adverse device effects. As a self-guided tool, *lipodia* provides a scalable and guideline-concordant means of delivering structured lifestyle support that is rarely available in routine care.

## Introduction

In Europe, cardiovascular disease (CVD) remains the leading contributor to premature death and to disability-adjusted life years^1^. Among its modifiable drivers, dyslipidaemia, an abnormal blood lipid profile most often marked by elevated low-density lipoprotein cholesterol (LDL-C), ranks among the most important: elevated cholesterol is present in about 39% of adults globally, and LDL-C ranked as the eighth-leading risk factor for death in 2019^2^. LDL-C is tied to cardiovascular risk in a causal, dose-dependent, and log-linear manner with no evident lower cut-off, so that each absolute drop in LDL-C yields a proportional fall in cardiovascular risk^3^.

Reducing LDL-C is accordingly the central goal of managing hypercholesterolaemia, the form of dyslipidaemia defined by elevated LDL-C. Across major guidelines, structured lifestyle modification is positioned as the foundation of lipid management, carrying a Class I recommendation across every risk category and LDL-C level^4–7^. Lifestyle change is not an optional adjunct but the first therapeutic step upon which pharmacological therapy may be added.

In everyday practice, however, this guideline-mandated basis is rarely provided. Dietary counselling takes place in only around 12% of physician encounters, and even high-risk patients receive it in roughly one in five cases, chiefly because of limited time, reimbursement, and training^8,9^. Thus, a substantial gap persists between guideline recommendations and the care most patients actually receive. Closing it calls for approaches that are not only effective but also widely accessible, engaging, and scalable, qualities that digital interventions may be particularly well suited to offer.

*lipodia* is a novel, comprehensive, fully self-guided digital health application for patients with hypercholesterolaemia. It incorporates established principles of effective behaviour-change support: it preserves patient autonomy, encourages specific goal-setting, tailors content to individual needs, and addresses lifestyle multifactorially rather than in isolated domains. In this pragmatic RCT we set out to determine whether *lipodia* lowers LDL-C and favourably affects other cardiometabolic risk factors when used under everyday, real-world conditions, compared to usual care (‘treatment as usual’; TAU) alone.

## Results

### Participant flow

Of 1,659 individuals screened, 278 met eligibility criteria and were randomised to *lipodia* (n = 140) or control (n = 138). Attrition was low and balanced: overall dropout (missing questionnaire or laboratory data) was 5.7% in the *lipodia* arm and 3.6% in the control arm (*χ²* = 0.293, *p* = .588); dropout defined by a missing primary-endpoint laboratory report at T2 (6 months following baseline) was 9.3% vs 5.8% (*χ²* = 0.763, *p* = .382). Participant flow is shown in Figure 1.

**Figure 1.**
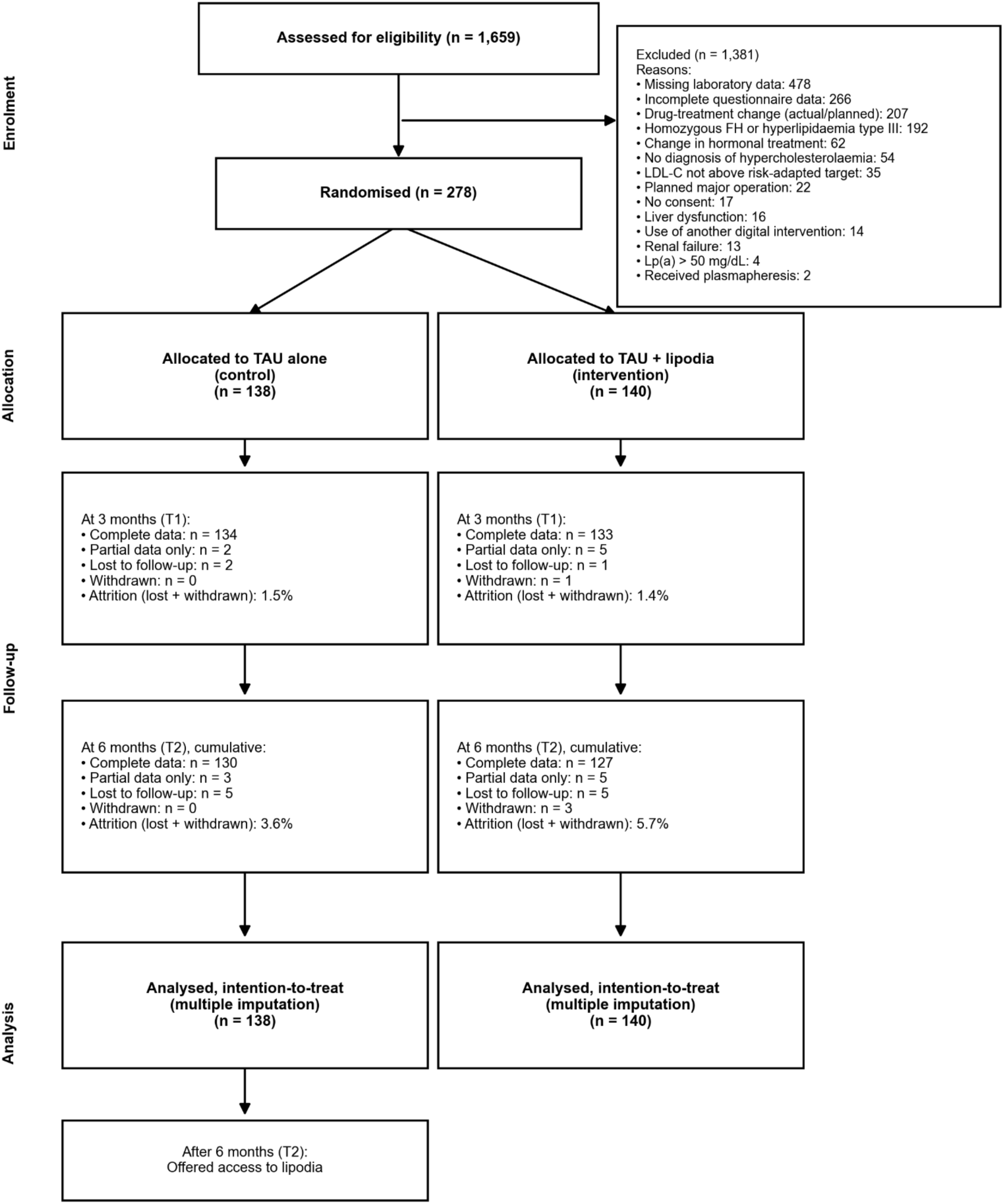
Flow of participants through the DIGICHOL trial. CONSORT diagram of enrolment, allocation, follow-up at 3 months (T1) and 6 months (T2), and analysis. Complete data denotes a completed questionnaire and a submitted laboratory report; partial data denotes that only one of the two was provided; lost to follow-up denotes that neither was provided. Attrition is the sum of participants lost to follow-up and withdrawn, shown as a percentage of the allocated arm. All randomised participants were included in the intention-to-treat analysis with multiple imputation. FH, familial hypercholesterolaemia; LDL-C, low-density lipoprotein cholesterol; Lp(a), lipoprotein(a); TAU, treatment as usual.

**Figure 2.**
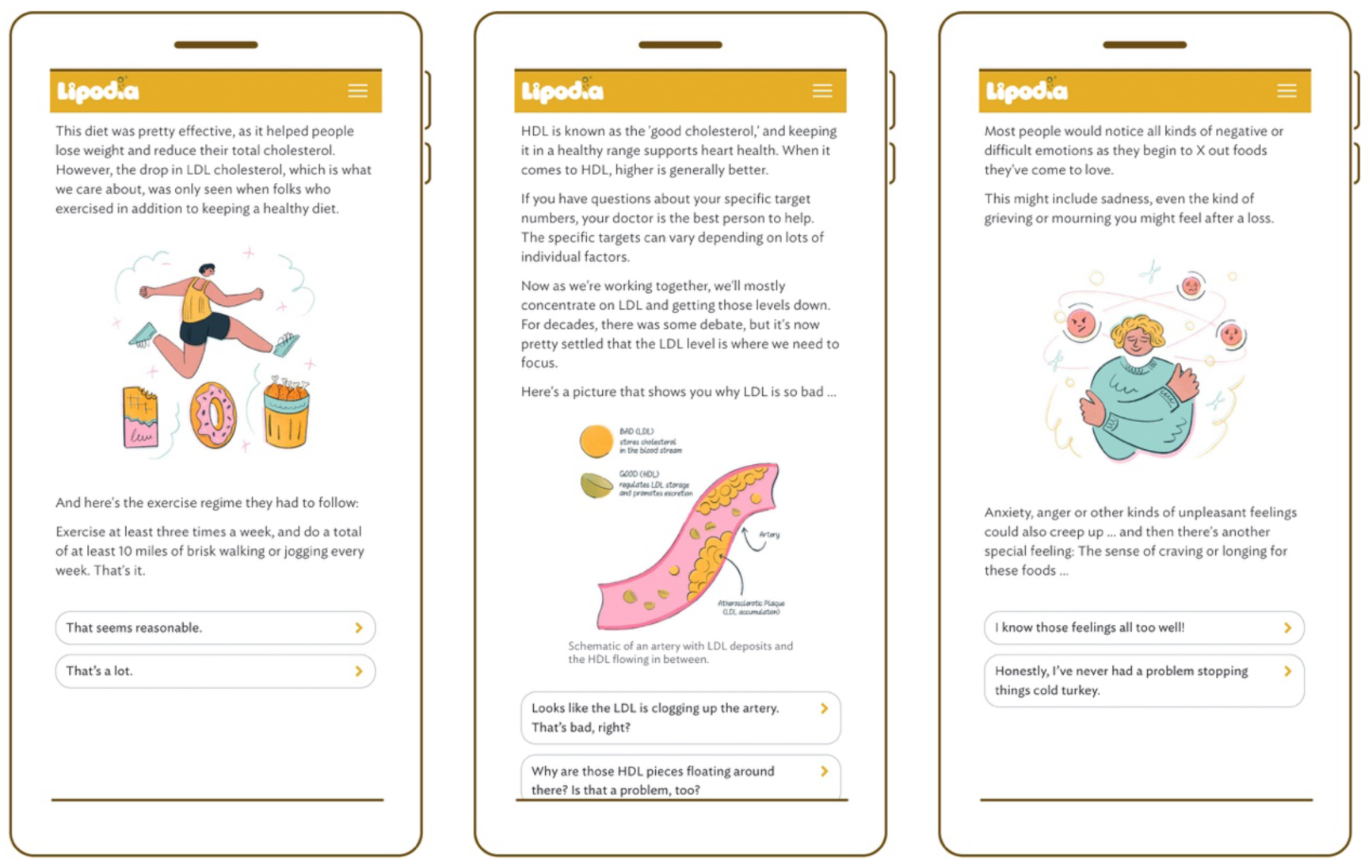
Screenshots from *lipodia*.

### Baseline characteristics

Baseline characteristics were comparable between groups (Table 1). Mean age was 57.2 years; 78.4% were women and 99.6% identified as White. Most participants were of low (42.4%) or moderate (30.9%) cardiovascular risk. Hypertension was the most common comorbidity (26.3%); 9.7% took any lipid-modifying medication and 6.5% took statins. Mean baseline LDL-C was 162.1 mg/dL.

**Table 1:** Baseline demographic and clinical characteristics.

| Characteristic | Control (n=138) | *lipodia* (n=140) | Total (n=278) |
| --- | --- | --- | --- |
| Age, years, mean (SD) | 57.7 (9.2) | 56.8 (10.2) | 57.2 (9.7) |
| Female, n (%) | 113 (81.9) | 105 (75.0) | 218 (78.4) |
| University degree, n (%) | 67 (48.6) | 77 (55.0) | 144 (51.8) |
| Hypertension, n (%) | 40 (29.0) | 33 (23.6) | 73 (26.3) |
| Diabetes, n (%) | 4 (2.9) | 2 (1.4) | 6 (2.2) |
| Any lipid-modifying medication (C10), n (%) | 14 (10.1) | 13 (9.3) | 27 (9.7) |
| Statin use, n (%) | 10 (7.2) | 8 (5.7) | 18 (6.5) |
| LDL-C, mg/dL, mean (SD) | 162.8 (37.0) | 161.5 (35.7) | 162.1 (36.3) |
| Non-HDL-C, mg/dL, mean (SD) | 178.7 (41.0) | 178.4 (39.1) | 178.5 (40.0) |
| Triglycerides, mg/dL, mean (SD) | 117.0 (56.5) | 119.7 (55.4) | 118.4 (55.8) |
| HDL-C, mg/dL, mean (SD) | 64.4 (17.9) | 62.2 (17.3) | 63.3 (17.6) |
| PAM-13, mean (SD) | 79.6 (11.7) | 80.7 (12.1) | 80.1 (11.9) |
| AQoL-8D, mean (SD) | 77.2 (10.1) | 78.3 (10.0) | 77.7 (10.1) |
| BMI, kg/m<sup>2</sup>, mean (SD) | 26.5 (6.1) | 26.3 (4.3) | 26.4 (5.3) |

*Note.* SD = standard deviation; LDL-C = low-density lipoprotein cholesterol; non-HDL-C = non-high-density lipoprotein cholesterol; HDL-C = high-density lipoprotein cholesterol; PAM-13 = Patient Activation Measure (13 items); AQoL-8D = Assessment of Quality of Life – 8 Dimensions; BMI = body mass index; C10 = ATC classification code for lipid-modifying agents.

### Primary endpoint: LDL-C at 6 months

The primary endpoint was met. In the ITT analysis, *lipodia* added to TAU produced a significantly greater relative reduction in LDL-C at 6 months than TAU alone: the baseline- and medication-adjusted between-group difference in %CfB was −3.9% (95% CI −7.4 to −0.4; p = .029; Cohen’s d = 0.26). Reductions in the lipodia group were approximately twofold greater than under TAU alone (−6.6% vs −2.9%). Under the conservative J2R sensitivity analysis, the point estimate remained essentially unchanged (−3.2%), with the p-value shifting marginally past the conventional cut-off (p = .054). The pre-specified analysis of post-treatment LDL-C scores is consistent (adjusted between-group difference −6.3 mg/dL; p = .046; Supplementary Table S1). Results of analyses for early effects at 3 months are reported in Table 2.

**Table 2:**
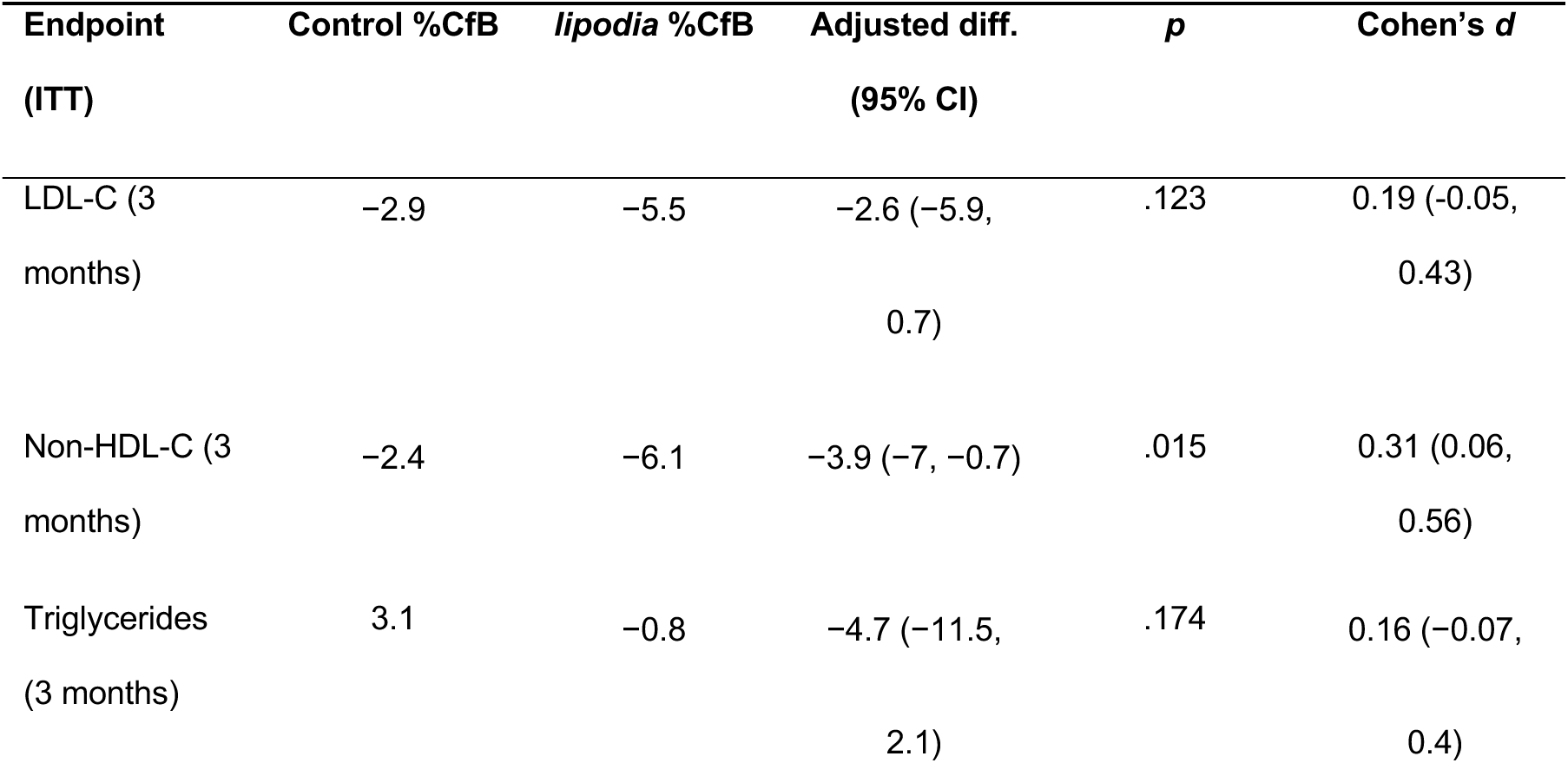
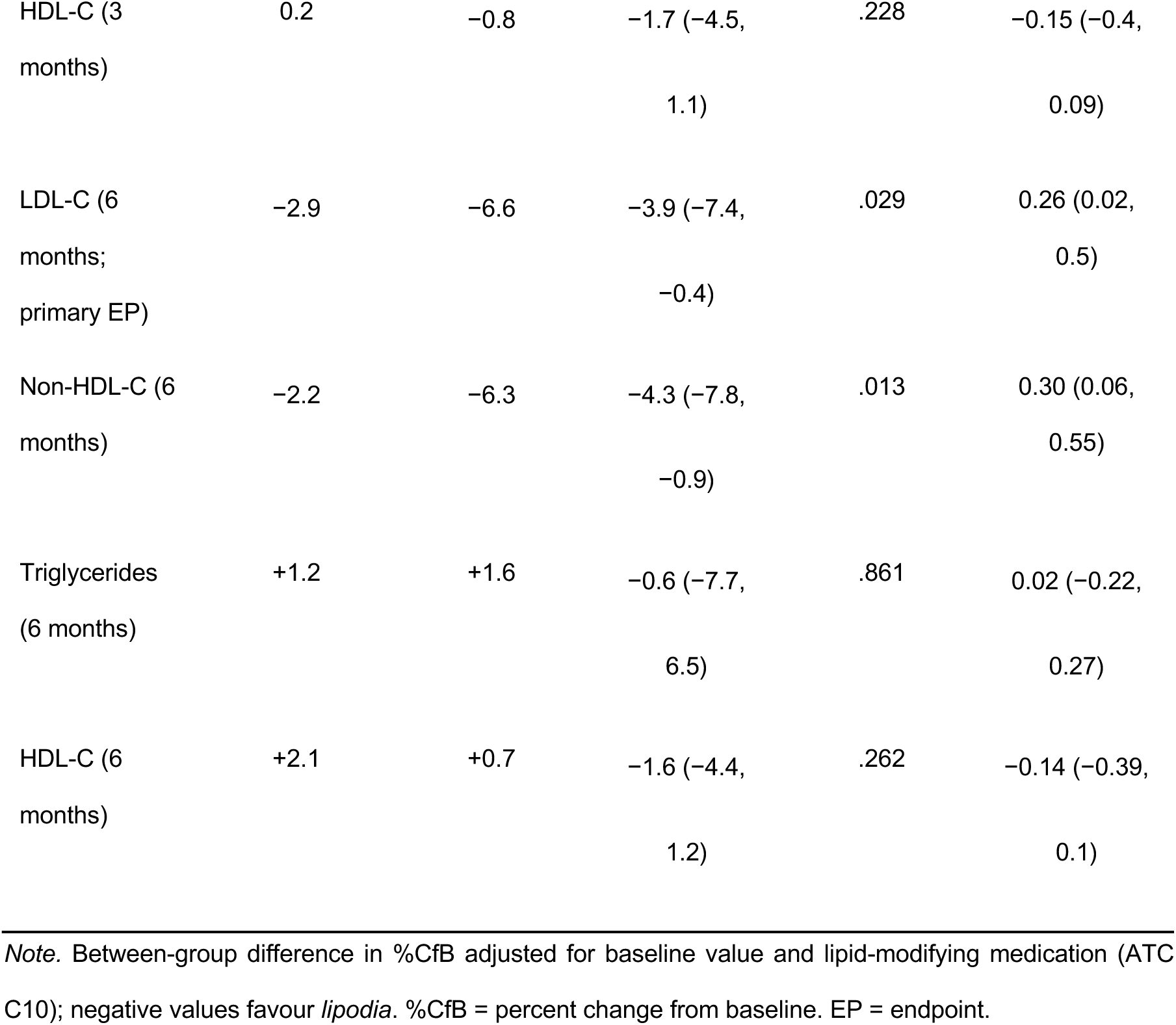
Relative change from baseline (%CfB) in lipid endpoints at 3 and 6 months (ITT, robust ANCOVA).

The pre-specified group-level MCID (a 10-percentage-point between-group difference in %CfB) was not met (3.9 points); this group-level MCID is, however, not an established criterion of individual clinical response. We therefore examined two within-person responder definitions. On a post-hoc but more established responder criterion (≥10% LDL-C reduction from baseline10), significantly more lipodia users responded than control participants (41.7% vs 23.8%; χ² = 9.34, p = .002). On the pre-specified criterion of attainment of the individual risk-adapted LDL-C target, a benchmark subject to strong floor effects, *lipodia* was favoured numerically but not significantly (11.0% vs 6.2%; *χ²* = 1.95, *p* = .163).

### Secondary endpoints

*lipodia* delivered significant, mutually reinforcing benefits across the confirmatory secondary endpoints tested in the gatekeeping sequence. At 6 months, patient activation improved significantly (adjusted PAM-13 difference +3.9 points, 95% CI 1.5 to 6.4; *p* = .002; d = 0.41), as did non-HDL-C (%CfB difference −4.3%, 95% CI −7.8 to −0.9; p = .013; d = 0.30) and health-related quality of life (adjusted AQoL-8D difference +3.2 points, 95% CI 1.6 to 4.7; p < .001; d = 0.48, the largest effect in the trial). No significant 6-month effect was observed for BMI (−0.6, 95% CI −1.3 to 0.1; p = .079; d = 0.24; point estimate favouring lipodia), triglycerides, or HDL-C. All three confirmatory secondary endpoints remained statistically significant under the conservative J2R analysis, and BMI became significant under J2R at 6 months (−0.8, p = .012). Intervention effects on patient activation, non-HDL-C, and quality of life were already significant at 3 months and consolidated by 6 months; see Table 2 for lipid endpoints and Table 3 for non-lipid endpoints.

**Table 3:** Post-treatment values and adjusted between-group differences for questionnaire/anthropometric secondary endpoints at 3 and 6 months (ITT, ANCOVA).

| Endpoint<br>(ITT) | Control | *lipodia* | Adjusted diff.<br>(95% CI) | *p* | Cohen's *d* |
| --- | --- | --- | --- | --- | --- |
| PAM-13<br>(points;<br>baseline) | 79.5 | 80.7 | - | - | - |
| AQoL-8D<br>(points;<br>baseline) | 77.2 | 78.2 | - | - | - |
| BMI (kg/m<sup>2</sup>;<br>baseline) | 26.5 | 26.2 | - | - | - |
| PAM-13<br>(points; 3<br>months) | 79.2 | 82.9 | +3 (0.8, 5.2) | .007 | 0.33 (0.1,<br>0.56) |
| AQoL-8D<br>(points; 3<br>months) | 76.4 | 79.3 | +2.1 (0.5, 3.6) | .009 | 0.32 (0.09,<br>0.56) |
| BMI (kg/m<sup>2</sup>; 3<br>months) | 26.3 | 25.7 | -0.4 (-0.6,<br>-0.2) | < .001 | 0.43 (0.16,<br>0.7) |
| PAM-13<br>(points; 6<br>months) | 79.3 | 84.0 | +3.9 (1.5, 6.4) | .002 | 0.41 (0.16,<br>0.66) |
| AQoL-8D<br>(points; 6<br>months) | 76.5 | 80.4 | +3.2 (1.6, 4.7) | <.001 | 0.48 (0.25,<br>0.7) |
| BMI (kg/m<sup>2</sup>; 6<br>months) | 26.3 | 25.6 | -0.6 (-1.3,<br>0.1) | .079 | 0.24 (-0.03,<br>0.5) |

Note. Values are baseline- and medication-adjusted between-group differences on the original scale; positive values for PAM-13/AQoL-8D and negative values for BMI favour *lipodia*. PAM-13 = Patient Activation Measure; AQoL-8D = Assessment of Quality of Life – 8 Dimensions; BMI = body mass index.

### Pre-specified subgroup analyses

The LDL-C treatment effect was concentrated in participants aged 18–65 (n = 227), who represented the large majority of the sample. In this subgroup, the adjusted %CfB difference was −4.5% (95% CI −8.4 to −0.5; p = .028; d = 0.30) and remained significant under J2R (−3.8%, p = .047); responder rates (≥10% reduction) were 42.7% vs 21.9% (χ² = 10.32, p = .001). The secondary endpoints (patient activation +3.6, p = .011; non-HDL-C −5.3%, p = .009; AQoL-8D +2.7, p = .003) were reproduced in this subgroup. In the smaller, less well-powered subgroup aged ≥66 (n = 51) the effect was not detectable. By sex, lipodia produced a significant LDL-C reduction in women (−4.2%, p = .026); in men both arms improved similarly, largely because about one in four control-group men newly initiated lipid-lowering medication during follow-up. The effect was directionally consistent across all cardiovascular risk categories (all favouring lipodia; largest in the high/very-high-risk stratum, d = 0.39), though no single stratum was individually significant given the reduced power of the split. It was reproduced in the statin-naïve majority (−3.8%, p = .036) and in those on no lipid-modifying medication (−3.9%, p = .026), while the small already-medicated subgroups were underpowered. Full subgroup results are provided in the Supplementary Material (Supplementary Table S5 and Supplementary Figure S1).

### Usage, satisfaction, and safety

Uptake was near-universal: 139 of 140 participants offered *lipodia* registered to use it (99.3%), and average weekly use was 38.4 minutes (SD 35.6). Satisfaction with the programme and with treatment was high (Net Promoter Score 29.2 at T1 and 33.3 at T2; ZUF-8 26.0 and 26.3 respectively), and patients reported subjective improvement in health and quality of life (PGIC > 4) at every timepoint. Adverse events (unplanned/emergency outpatient or inpatient treatment) occurred at comparably low rates in both arms (T1: 7.3% *lipodia* vs 10.3% control, *p* = .382; T2: 5.3% vs 7.5%, *p* = .472). None of the adverse events were attributed to *lipodia*, and there were no adverse device effects, device deficiencies, serious adverse events, or deaths.

## Discussion

In this pragmatic RCT, the self-guided digital application *lipodia* added to usual care significantly reduced LDL-C at 6 months relative to usual care alone. Additionally, non-HDL-C fell, patient activation rose, and health-related quality of life improved, with all three between-group differences reaching statistical significance. Together, these findings indicate a broad and consistent pattern of benefit. Under a deliberately conservative jump-to-reference sensitivity analysis, the primary effect was directionally stable, with a virtually unchanged point estimate (−3.2% vs. −3.9%), although the p-value shifted just above the conventional threshold (p = .054); key secondary effects, most notably non-HDL-C, remained significant under the same assumptions, and benefits emerged early (3 months) and consolidated by 6 months. Taken together, this convergence across endpoints, timepoints, and analyses points to the robust and holistic effect of *lipodia* in helping people with dyslipidaemia make clinically meaningful lifestyle changes.

The pattern of effects is the coherent signature of a single mechanism: *lipodia* works not through pharmacology but by building patients’ knowledge, skills, and confidence to take charge of their own health (i.e. patient activation), which then drives the behaviours that produce downstream physiological benefit. Given that LDL-C relates to cardiovascular risk causally, dose-dependently, and without a lower threshold, the LDL-C reduction produced by *lipodia* maps directly onto reduced cardiovascular risk. Importantly, the effect should be judged against other fully self-guided digital interventions, for which meta-analyses report average between-group LDL-C reductions on the order of 4–8 mg/dL relative to usual care (often non-significant)^11,12^, rather than against the idealised maximal efficacy of statins, which most patients never realise, given real-world adherence below 60%^13^. We do not claim that *lipodia* matches the magnitude of optimally taken pharmacotherapy, and it is not intended to. Its value lies in a fundamentally different risk-benefit profile: a significant, guideline-concordant LDL-C reduction achieved with no pharmacological side effects, near-limitless scalability, and high uptake with low attrition. Just as importantly, user and treatment satisfaction were consistently high, indicating that patients not only used *lipodia* but valued the experience. Unlike lipid-lowering drugs, lipodia additionally improved patient activation and health-related quality of life, patient-relevant outcomes that pharmacotherapy does not deliver. For the large group of patients who are unwilling to start a statin, are intolerant of one, or remain above target despite treatment, this combination makes lipodia a clinically meaningful complement rather than a weaker substitute.

Although the pre-specified group-level MCID (a 10-percentage-point between-group difference in %CfB) was not reached, such a group-level threshold is not a recognised marker of individual clinical response and imposed a demanding, drug-level standard on a self-guided lifestyle programme added to active usual care. Clinical importance is better judged at the level of the individual: using the within-person criterion (a ≥10% LDL-C reduction10), lipodia roughly doubled the share of responders compared with usual care. Taken together with the significant primary endpoint and the concordant improvements across atherogenic lipids, patient activation, and quality of life, this individual-level gain is the more appropriate gauge of the intervention’s value.

The trial has several notable strengths. Its pragmatic, fully online design mirrors the conditions under which a self-guided application would actually be used, so the estimates speak to real-world effectiveness rather than to efficacy under idealised conditions. The primary endpoint was an objective, laboratory-measured lipid concentration, which is immune to the expectancy effects that can inflate self-reported outcomes in unblinded behavioural trials. Type I error across the family of endpoints was controlled through a pre-specified gatekeeping strategy, and the conservative jump-to-reference sensitivity analyses guarded against optimistic assumptions about missing data. Finally, uptake was near-universal and attrition was low and balanced between arms, which limits the scope for attrition bias and strengthens the internal validity of the comparison.

Several limitations should be considered, all characteristic of pragmatic trials of self-guided digital interventions. The sample was predominantly female and highly educated, mirroring the real-world user base of digital health tools and thereby supporting rather than limiting the relevance of the findings for that population. Laboratory values were obtained from participants’ own certified laboratories rather than a central laboratory; any resulting variability is non-differential and, if anything, biases towards the null. Participants and staff could not be blinded, but the objective primary endpoint makes the primary result robust to this, and self-report is the appropriate measurement for patient-reported outcomes such as quality of life.

Added to usual care, *lipodia* delivered a significant and clinically coherent constellation of benefits spanning atherogenic lipids, patient activation, and health-related quality of life, alongside an excellent safety and acceptance profile. Being fully self-guided, it provides a scalable and guideline-concordant means of supplying the structured lifestyle basis of lipid management that the guidelines uniformly recommend, yet routine care rarely delivers, helping patients lower their LDL-C and, in turn, their cardiovascular risk.

## Methods

### Trial design

DIGICHOL was a prospective, two-arm, parallel-group, randomised (1:1) controlled trial conducted entirely online, without a traditional physical investigation site. The pragmatic design was chosen to evaluate effectiveness in a routine, real-world setting. The trial was conducted in accordance with ISO 14155 and the ethical principles of the Declaration of Helsinki, was approved by the Ethics Committee of the Medical Faculty of the Christian-Albrechts-Universität zu Kiel (D559/24), and was conducted according to a predefined Clinical Investigation Plan (CIP) prepared in accordance with ISO 14155; the CIP included the full statistical analysis plan. The CIP is available from the corresponding author on reasonable request. Participants were enrolled between 26 January and 12 November 2025, with follow-up continuing until the last 6-month visit (T2) on 22 May 2026.

### Participants

Recruitment used targeted online advertising that directed interested individuals to a dedicated study website. Eligible participants were adults (≥18 years) with a physician-and laboratory-confirmed diagnosis of hypercholesterolaemia (ICD-10 E78.0, E78.2, E78.4, E78.5, E78.8, E78.9), LDL-C above their risk-adapted target, triglycerides <400 mg/dL, stable drug treatment (last 4 weeks) and hormonal treatment (last 6 months), who had been made aware of lifestyle measures by a physician, provided informed consent, and had sufficient German-language skills. Key exclusion criteria were homozygous familial hypercholesterolaemia, hyperlipidaemia type III, plasmapheresis, Lp(a) >50 mg/dL, current/planned pregnancy, planned major surgery, liver dysfunction, end-stage renal failure, planned changes to (hormonal) drug treatment within 6 months, and prior use of another digital intervention for dyslipidaemia. All consent and data collection were online.

### Randomisation and allocation concealment

A randomisation list was created before enrolment with a Python script that drew computer-generated random numbers and applied block randomisation (1:1; variable blocks of 4, 8, or 16). The list was embedded in the password-protected study platform (read-only and immutable after activation) with role-based access restricted to the study manager, who had no participant contact before randomisation. Participants received an automated e-mail with their allocation, ensuring full allocation concealment. Owing to the nature of the intervention, participants and study staff could not be blinded to allocation; the primary endpoint was an objective, laboratory-measured biomarker, which is robust to the absence of participant blinding. Data analysts were not blinded to group allocation.

### Intervention (*lipodia*)

*lipodia* is an interactive, internet-based, fully self-guided programme that provides psychoeducational content grounded in evidence-based psychological and psychotherapeutic methods to support lifestyle change in people with hypercholesterolaemia. Content is delivered through simulated dialogues in which users react to information by selecting predefined responses, and the programme adapts content accordingly. A core set of behavioural lipid-management modules (e.g. dietary habits, physical activity, willpower/impulse control) is combined with optional modules fostering autonomy and individualisation (stress, mood, sleep, and weight management; quitting smoking and drinking). Optional daily short text messages provide reminders and motivation. *lipodia* requires only internet access and a web browser (no installation) and is intended as an adjunct to, not a substitute for, usual care.

### Control group and concomitant treatment

Participants in the control arm continued their usual medical care in discussion with their treating team. Consistent with the pragmatic design, usual care reflected the reality of routine care and could comprise primary-care, specialist treatment, psychotherapy, or no treatment. *lipodia* was provided free of charge; intervention participants received immediate access after randomisation, and control participants were offered access after 6 months.

### Outcomes

The primary endpoint was the relative change from baseline (%CfB) in fasting LDL-C at 6 months (T2). Pre-specified secondary endpoints, tested in a fixed gatekeeping sequence, were: patient activation (PAM-13, German version^14^); %CfB in fasting non-HDL-C; health-related quality of life (AQoL-8D, German version^15^); BMI; %CfB in fasting triglycerides; %CfB in fasting HDL-C; and %CfB in LDL-C at T1 (3 months). Corresponding 3-month assessments of the other outcomes were exploratory. The outcomes were selected for their relevance to decision-makers: LDL-C is a causal, guideline-endorsed treatment target, while patient activation and quality of life capture patient-relevant benefits not provided by pharmacotherapy. The six-month horizon is long enough to demonstrate a clinically interpretable lipid change and matches the timeframe over which a self-guided tool would be assessed in routine care. Adherence, user satisfaction (Net Promoter Score^16^, ZUF-8^17^, PGIC^18^), and adverse events / adverse device effects were also assessed. At each timepoint, both an online questionnaire and a laboratory report (from a certified laboratory in geographical proximity of the participant, kept constant across timepoints) were required.

### Sample size

The sample size was driven by the primary endpoint. Assuming a 10-percentage-point between-group difference in mean percent LDL-C reduction, 90% power (1−β), and a two-sided α of 0.05, an analysis of variance required 226 participants. After allowing for 20% attrition by 6 months, the planned enrolment was 272 participants (1:1); ultimately, 278 were randomised.

### Statistical analysis

Analyses were conducted in R version 4.6.0. Effectiveness at T2 was analysed by ANCOVA comparing groups on %CfB (for lipid endpoints) or post-treatment values (PAM-13, AQoL-8D, BMI), adjusting for the baseline value and intake of lipid-modifying medication (ATC C10). Because the normality assumption was violated for %CfB outcomes, a robust implementation of ANCOVA was used. Standardised effect sizes (Cohen’s *d*) were derived from the adjusted between-group difference. The primary analysis followed the intention-to-treat (ITT) principle with bootstrapped maximum-likelihood multiple imputation under a missing-at-random (MAR) assumption (bootImpute^19^ and mice^20^ packages). Imputation models included baseline scores, group allocation, and additional sociodemographic and clinical variables (age, sex, diabetes, hypertension, BMI, intake of lipid-modifying agents (ATC code C10) at baseline). A jump-to-reference (J2R) imputation served as a conservative sensitivity analysis (bootImpute and mlmi^19^ packages). A pre-specified gatekeeping strategy controlled the family-wise error rate, so no further multiplicity adjustment was required; all tests used a two-sided 5% significance level. A group-level minimal clinically important difference (MCID) was pre-specified as a 10-percentage-point between-group difference in %CfB LDL-C. Additionally, attainment of the risk-adapted LDL-C target was prespecified as a within-person responder analysis. We additionally conducted a post-hoc within-person responder analysis regarding % CfB (≥10% LDL-C reduction from baseline10), because a group-level MCID for % CfB is not an established responder criterion. Both individual-level responder analyses were conducted on complete-case data and compared between groups using *χ²* tests. Pre-specified subgroup analyses (including age and sex) were performed for the primary endpoint. Note that the original study protocol planned to compare post-treatment values for all lipid and non-lipid endpoints. Following the advice of a regulatory body, and in line with the applicable EMA guidance (EMA/CHMP/748108/2013 Rev. 3^21^), the primary analysis for the lipid endpoints was changed to %CfB in place of post-treatment scores. The corresponding pre-specified analyses of post-treatment scores for all lipid endpoints are provided in the Supplementary Material (Supplementary Tables S1–S4).

### Patient and public involvement

Patients and members of the public were not formally involved in the design, conduct, or reporting of this trial. The intervention itself was developed with input from prospective users during earlier formative work.

### Use of large language models

A large language model (Claude Opus 4.8) was used to assist with language editing and formatting of this manuscript draft. The authors take full responsibility for the content, the accuracy of all reported data, and the interpretation of the results.

## Data availability

The dataset analysed here is not publicly available because participants did not consent to open data sharing and the data contain potentially identifying health information. De-identified individual participant data underlying the results, together with the data dictionary, can be shared with researchers who provide a methodologically sound proposal, subject to approval by the sponsor (GAIA) and a signed data-sharing and data-protection agreement. Proposals should be directed to the corresponding author (Antje Riepenhausen) and can be submitted up to 36 months after publication of this Article.

## Code availability

The statistical analysis code used to generate the results reported here is available from the corresponding author on the same terms and within the same time window as the data.

## Acknowledgements

The authors would like to thank all participants for their valuable contribution to this study. This trial was funded by GAIA, the manufacturer and developer of *lipodia*. GAIA provided the lipodia application free of charge and contributed to the trial design, the analysis and interpretation of the data, and the preparation of the manuscript through authors employed by the company (see Author contributions and Competing interests). The decision to submit the manuscript for publication was made jointly by all authors.

## Author contributions

AR: conceptualisation; methodology; data curation; formal analysis; visualisation; writing - original draft; writing - review and editing. CE: data curation; formal analysis; visualisation; writing - review and editing. BM: conceptualisation; methodology. KJC: conceptualisation; methodology; writing - review and editing; supervision.

## Competing interests

AR, CE, and BM are employees of GAIA, the developer, owner, and distributor of *lipodia*. KJC declares no competing interests.

## Ethics approval and trial registration

The trial was approved by the Ethics Committee of the Medical Faculty of the Christian-Albrechts-Universität zu Kiel (D559/24), conducted in accordance with ISO 14155 and the Declaration of Helsinki, and registered at ClinicalTrials.gov (NCT05988866; https://clinicaltrials.gov/study/NCT05988866?term=NCT05988866&viewType=Card&rank=1; first posted 2023-08-14). All participants provided informed consent online.

## Notes

### Clinical Trial

NCT05988866

### Author Declarations

Ethics committee of Christian-Albrechts-Universität zu Kiel, Germany gave ethical approval for this work (D559/24)

