## Supplementary Material for "A self-guided digital health application for LDL-cholesterol reduction in adults with hypercholesterolaemia (The DIGICHOL randomised controlled trial)"

This supplement provides additional pre-specified analyses that complement the main text. The first part (Supplementary Tables S1 to S4) reports the pre-specified analyses of post-treatment scores for the four lipid endpoints (LDL-C, non-HDL-C, triglycerides, and HDL-C). The second part (Supplementary Table S5 and Supplementary Figure S1) reports the pre-specified subgroup analyses for the primary endpoint (LDL-C). All analyses follow the intention-to-treat principle with multiple imputation under a missing-at-random assumption and are adjusted for the baseline value and lipid-modifying medication (ATC C10), as in the main analysis. The third part includes the CONSORT 2025 checklist.

#### 1. Post-treatment-score analyses (lipid endpoints)

As pre-registered, each lipid endpoint was originally analysed as the post-treatment concentration with adjustment for the baseline value; following regulatory advice and applicable EMA guidance, the main text instead reports the relative change from baseline (%CfB). The tables below give the originally pre-specified post-treatment-score analyses at 3 months (T1) and 6 months (T2), with baseline (T0) values for reference.

**Supplementary Table S1**

*LDL-C (mg/dL): pre-specified analysis of post-treatment scores (ITT, baseline-adjusted ANCOVA).*

| Timepoint | Control, mean (SD) | lipodia, mean (SD) | Adjusted difference (95% CI) | p | Cohen’s d (95% CI) |
|---|---|---|---|---|---|
| Baseline (T0) | 162.7 (36.3) | 161.6 (35.5) | — | — | — |
| 3 months (T1) | 156.1 (38.8) | 151.7 (37.3) | −3.5 (−9.7, 2.7) | .265 | 0.14 (−0.10, 0.38) |
| 6 months (T2) | 157.2 (41.3) | 149.9 (38.1) | −6.3 (−12.5, −0.1) | .046 | 0.24 (0.00, 0.48) |

*Note.* Values are post-treatment concentrations in mg/dL. The adjusted between-group difference is the baseline- and medication-adjusted (ATC C10) difference in post-treatment score (lipodia minus control) from an ITT ANCOVA with multiple imputation under a missing-at-random assumption (n = 138 control, n = 140 lipodia). For LDL-C, non-HDL-C, and triglycerides a negative difference favours lipodia; for HDL-C higher values are favourable, so a negative difference does not favour lipodia. In the main text these lipid endpoints are reported as relative change from baseline (%CfB).

#### Supplementary Table S2

*Non-HDL-C (mg/dL): pre-specified analysis of post-treatment scores (ITT, baseline-adjusted ANCOVA).*

| Timepoint | Control, mean (SD) | lipodia, mean (SD) | Adjusted difference (95% CI) | p | Cohen's d (95% CI) |
|---|---|---|---|---|---|
| Baseline (T0) | 178.6 (40.2) | 178.5 (38.8) | — | — | — |
| 3 months (T1) | 172.4 (42.3) | 166.7 (39.6) | −5.6 (−12.2, 0.9) | .093 | 0.21 (−0.04, 0.46) |
| 6 months (T2) | 173.9 (46.0) | 166.3 (41.7) | −7.4 (−14.1, −0.7) | .030 | 0.26 (0.02, 0.50) |

*Note.* Values are post-treatment concentrations in mg/dL. The adjusted between-group difference is the baseline- and medication-adjusted (ATC C10) difference in post-treatment score (lipodia minus control) from an ITT ANCOVA with multiple imputation under a missing-at-random assumption (n = 138 control, n = 140 lipodia). For LDL-C, non-HDL-C, and triglycerides a negative difference favours lipodia; for HDL-C higher values are favourable, so a negative difference does not favour lipodia. In the main text these lipid endpoints are reported as relative change from baseline (%CfB).

#### Supplementary Table S3

*Triglycerides (mg/dL): pre-specified analysis of post-treatment scores (ITT, baseline-adjusted ANCOVA).*

| Timepoint | Control, mean (SD) | lipodia, mean (SD) | Adjusted difference (95% CI) | p | Cohen's d (95% CI) |
|---|---|---|---|---|---|
| Baseline (T0) | 117.0 (56.1) | 119.6 (55.0) | — | — | — |
| 3 months (T1) | 116.4 (58.9) | 111.9 (51.3) | −6.3 (−15.2, 2.6) | .167 | 0.17 (−0.07, 0.40) |

| | | | | | |
|---|---|---|---|---|---|
| 6 months (T2) | 113.0 (55.0) | 114.2 (53.1) | −0.6 (−10.3, 9.2) | .908 | 0.01 (−0.23, 0.26) |

*Note.* Values are post-treatment concentrations in mg/dL. The adjusted between-group difference is the baseline- and medication-adjusted (ATC C10) difference in post-treatment score (lipodia minus control) from an ITT ANCOVA with multiple imputation under a missing-at-random assumption (n = 138 control, n = 140 lipodia). For LDL-C, non-HDL-C, and triglycerides a negative difference favours lipodia; for HDL-C higher values are favourable, so a negative difference does not favour lipodia. In the main text these lipid endpoints are reported as relative change from baseline (%CfB).

**Supplementary Table S4**

*HDL-C (mg/dL): pre-specified analysis of post-treatment scores (ITT, baseline-adjusted ANCOVA).*

| Timepoint | Control, mean (SD) | lipodia, mean (SD) | Adjusted difference (95% CI) | p | Cohen's d (95% CI) |
|---|---|---|---|---|---|
| Baseline (T0) | 64.3 (17.8) | 62.2 (17.1) | — | — | — |
| 3 months (T1) | 63.7 (17.4) | 61.3 (16.9) | −0.5 (−2.4, 1.4) | .611 | −0.06 (−0.30, 0.17) |
| 6 months (T2) | 65.1 (17.5) | 61.7 (15.3) | −1.5 (−3.4, 0.3) | .108 | −0.20 (−0.44, 0.04) |

*Note.* Values are post-treatment concentrations in mg/dL. The adjusted between-group difference is the baseline- and medication-adjusted (ATC C10) difference in post-treatment score (lipodia minus control) from an ITT ANCOVA with multiple imputation under a missing-at-random assumption (n = 138 control, n = 140 lipodia). For LDL-C, non-HDL-C, and triglycerides a negative difference favours lipodia; for HDL-C higher values are favourable, so a negative difference does not favour lipodia. In the main text these lipid endpoints are reported as relative change from baseline (%CfB).

### 2. Subgroup analyses (primary endpoint, LDL-C)

The following pre-specified subgroup analyses examine whether the LDL-C treatment effect at 6 months (T2) differed across clinically relevant strata. Estimates are the relative change from baseline (%CfB) from robust ANCOVA. The results are summarised numerically in Supplementary Table S5 and graphically in Supplementary Figure S1.

**Supplementary Table S5**

*Pre-specified LDL-C subgroup analyses at 6 months (T2, %CfB, robust ANCOVA).*

| Subgroup | Difference in %CfB (95% CI) | Cohen's d (95% CI) | p |
|---|---|---|---|
| ***Age*** | | | |
| 18-65 years (n = 227) | −4.5 (−8.4, −0.5) | 0.30 (0.02, 0.57) | .028 |
| ≥ 66 years (n = 51) | −0.1 (−8.2, 8.0) | 0.03 (−0.59, 0.64) | .978 |
| ***Sex*** | | | |
| Women (n = 218) | −4.2 (−7.9, −0.5) | 0.31 (0.03, 0.58) | .026 |
| Men (n = 60) | +2.1 (−9.3, 13.6) | −0.09 (−0.69, 0.51) | .714 |
| ***Cardiovascular risk category*** | | | |
| Low (n = 118) | −5.1 (−11.7, 1.6) | 0.32 (−0.11, 0.75) | .135 |
| Moderate (n = 86) | −1.9 (−8.4, 4.5) | 0.15 (−0.32, 0.61) | .554 |
| High / very high (n = 74) | −5.0 (−11.1, 1.1) | 0.39 (−0.10, 0.89) | .109 |
| ***Any lipid-modifying medication at baseline (ATC C10)*** | | | |
| On C10 medication (n = 27) | −2.6 (−23.9, 18.6) | 0.13 (−0.83, 1.09) | .807 |
| Not on C10 medication (n = 251) | −3.9 (−7.4, −0.5) | 0.28 (0.02, 0.54) | .026 |
| ***Statin at baseline (ATC C10AA)*** | | | |
| Not on a statin (n = 260) | −3.8 (−7.3, −0.3) | 0.26 (0.01, 0.52) | .036 |
| ***Change in regular C10 medication during follow-up*** | | | |
| Changed (n = 26) | −1.7 (−46.3, 42.9) | 0.35 (−21.04, 21.73) | .941 |

| | | | |
|---|---|---|---|
| No change (n = 239) | −4.2 (−7.4, −1.0) | 0.32 (0.07, 0.58) | .010 |

*Note.* Pre-specified subgroup analyses for the primary endpoint (LDL-C) at 6 months, expressed as relative change from baseline (%CfB) from a robust ANCOVA adjusted for baseline value and lipid-modifying medication (ATC C10). The between-group difference is lipodia minus control; a negative value favours lipodia. Cohen's d and its 95% confidence interval are the values shown in Supplementary Figure S1. The on-statin stratum (n = 18) was too small for a stable ANCOVA and is therefore not reported. Cardiovascular risk categories follow the ESC/EAS classification. Subgroup analyses were not powered for interaction tests and are interpreted as exploratory.

### Supplementary Figure S1

*Pre-specified LDL-C subgroup analyses at 6 months (T2): forest plot of standardised effects (Cohen's d).*

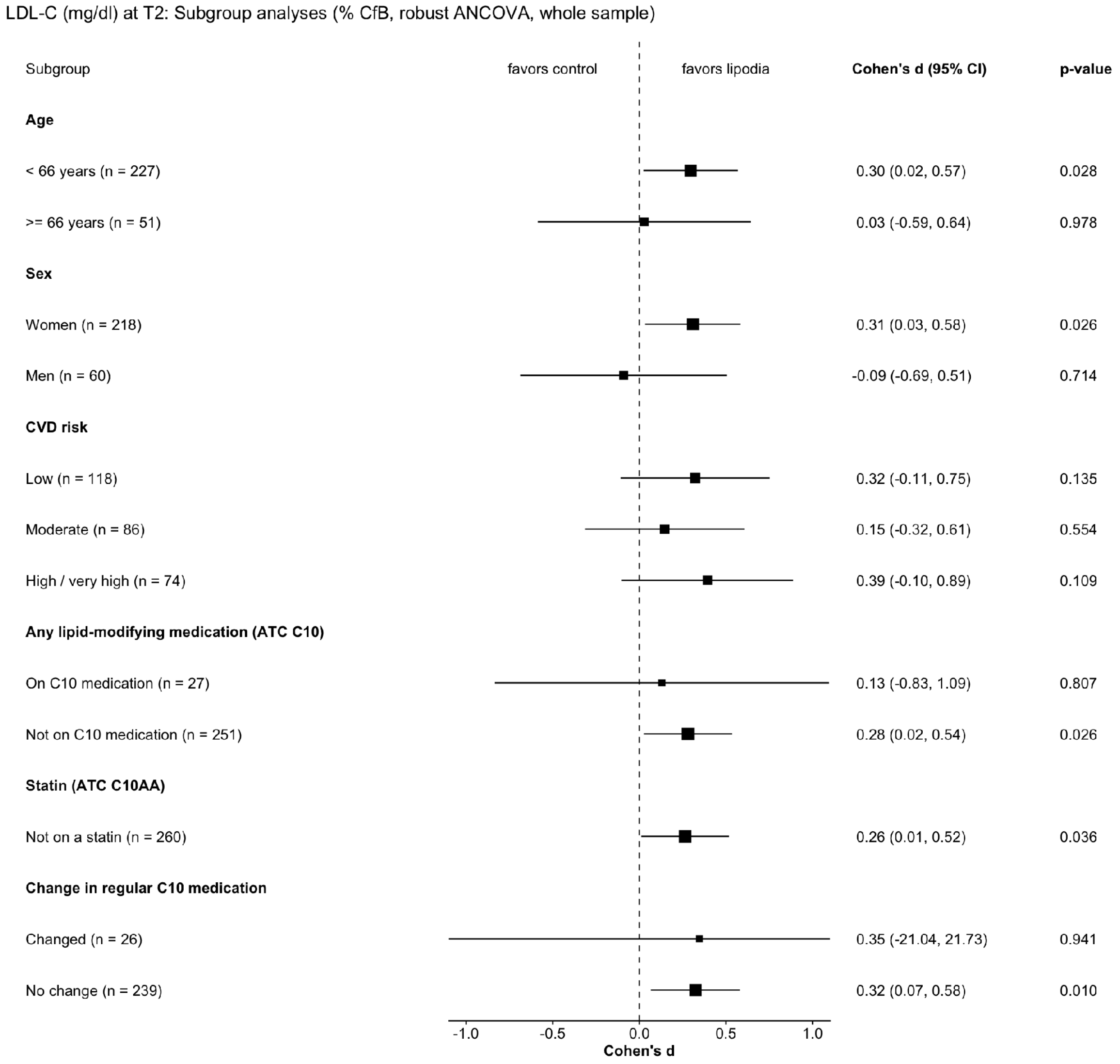

*Note.* Forest plot of the pre-specified LDL-C subgroup analyses at 6 months (T2), expressed as Cohen's d (standardised between-group effect on %CfB) with 95% confidence intervals from robust ANCOVA. Points to the right of the dashed line favour lipodia. Numeric estimates and p-values are given in Supplementary Table S5.

### 3. CONSORT 2025 checklist

| Section/Topic | Item No | CONSORT 2025 checklist item | Reported in manuscript |
|---|---|---|---|
| **Title and abstract** | | | |
| | 1a | Identification as a randomised trial in the title | Subtitle: "The DIGICHOL randomised controlled trial"; Abstract *(Manuscript: lines 1–3 (title/subtitle); abstract lines 17–29)* |
| | 1b | Structured summary of the trial design, methods, results, and conclusions | Abstract (unstructured per npj Digital Medicine format; design, methods, results and conclusions are all covered) *(Manuscript: lines 17–29)* |
| **Open science** | | | |
| **Trial registration** | 2 | Name of trial registry, identifying number (with URL) and date of registration | Ethics approval and trial registration (ClinicalTrials.gov NCT05988866, with full study URL and first-posted date 2023-08-14) *(Manuscript: lines 12 and 371–376 (URL and first-posted date at lines 374–376))* |
| **Protocol and statistical analysis plan** | 3 | Where the trial protocol and statistical analysis plan can be accessed | Methods – Trial design (the statistical analysis plan is contained within the ISO 14155 Clinical Investigation Plan (CIP), which is available from the corresponding author on reasonable request) *(Manuscript: lines 229–239)* |
| **Data sharing** | 4 | Where and how the individual de-identified participant data (including data dictionary), statistical code and any other materials can be accessed | Data availability and Code availability statements (de-identified individual participant data plus data dictionary shareable on a methodologically sound proposal, subject to sponsor (GAIA) approval and a signed data-sharing/data-protection agreement, directed to the corresponding author, up to 36 months after publication; statistical code available on the same terms) *(Manuscript: lines 344–352 (data) and 353–355 (code))* |
| **Funding** | 5a | Sources of funding and other support (e.g. supply of drugs), and role of funders in the design, | Acknowledgements/Funding (lipodia developed and funded by GAIA; the funder's role in trial design, analysis, interpretation, and |

| | | | |
|---|---|---|---|
| | | conduct, analysis and reporting of the trial | manuscript preparation through employed authors is stated, and the decision to submit was made jointly by all authors) *(Manuscript: lines 356–362)* |
| **Conflicts of interest** | 5b | Financial and other conflicts of interest of the manuscript authors | Competing interests (several authors are employees of GAIA AG, the manufacturer of lipodia) *(Manuscript: lines 368–370)* |
| **Introduction** | | | |
| **Background and rationale** | 6 | Scientific background and rationale | Introduction (paragraphs 1–3) *(Manuscript: lines 31–51)* |
| **Objectives** | 7 | Specific objectives related to benefits and harms | Introduction (final paragraph: aim of the trial) *(Manuscript: lines 56–58)* |
| **Methods** | | | |
| **Patient and public involvement** | 8 | Details of patient or public involvement in the design, conduct and reporting of the trial | Methods – Patient and public involvement (patients and the public were not formally involved; the intervention was developed with input from prospective users during earlier formative work) *(Manuscript: lines 336–339)* |
| **Trial design** | 9 | Description of trial design including type of trial (e.g. parallel group, crossover), allocation ratio, and framework (e.g. superiority, equivalence, non-inferiority, exploratory) | Methods – Trial design (two-arm, parallel-group, 1:1, superiority; fully online) *(Manuscript: lines 230–232)* |
| **Changes to trial protocol** | 10 | Important changes to the trial after it commenced including any outcomes or analyses that were not prespecified, with reason | Methods – Statistical analysis (%CfB reported as the primary analysis of the lipid endpoints in place of post-treatment scores, following regulatory advice and applicable EMA guidance; the within-person 10% responder criterion is flagged as post hoc) *(Manuscript: lines 324–333)* |
| **Trial setting** | 11 | Settings (e.g. community, hospital) and locations (e.g. countries, sites) where the trial was conducted | Methods – Trial design and Participants (fully online; Germany) *(Manuscript: line 231; lines 240–252)* |
| **Eligibility criteria** | 12a | Eligibility criteria for participants | Methods – Participants *(Manuscript: lines 241–252)* |
| | 12b | If applicable, eligibility criteria for sites and for individuals delivering the interventions (e.g. | Not applicable – self-guided digital intervention; no sites or providers deliver the intervention |

| | | | |
|---|---|---|---|
| | | surgeons, physiotherapists) | |
| **Intervention and comparator** | 13 | Intervention and comparator with sufficient details to allow replication. If relevant, where additional materials describing the intervention and comparator (e.g. intervention manual) can be accessed | Methods – Intervention (lipodia); Control group and concomitant treatment [consider linking screenshots or a module flowchart in the Supplementary Information] *(Manuscript: lines 263–276 and 277–283)* |
| **Outcomes** | 14 | Prespecified primary and secondary outcomes, including the specific measurement variable, analysis metric, method of aggregation, and time point for each outcome | Methods – Outcomes (LDL-C relative change from baseline at 6 months as primary; secondary lipids, PAM-13, AQoL-8D at T1 and T2) *(Manuscript: lines 285–299)* |
| **Harms** | 15 | How harms were defined and assessed (e.g. systematically, non-systematically) | Methods – Outcomes/Safety (definition and assessment of adverse events and adverse device effects) *(Manuscript: lines 295–296 (Methods); lines 157–159 (Results))* |
| **Sample size** | 16a | How sample size was determined, including all assumptions supporting the sample size calculation | Methods – Sample size *(Manuscript: lines 301–305)* |
| | 16b | Explanation of any interim analyses and stopping guidelines | Not applicable – no interim analyses or stopping rules were used |
| **Randomisation: sequence generation** | 17a | Who generated the random allocation sequence and the method used | Methods – Randomisation and allocation concealment (computer-generated sequence) *(Manuscript: lines 254–256)* |
| | 17b | Type of randomisation and details of any restriction (e.g. stratification, blocking and block size) | Methods – Randomisation (block randomisation; variable blocks of 4, 8, or 16) *(Manuscript: lines 255–256)* |
| **Allocation concealment mechanism** | 18 | Mechanism used to implement the random allocation sequence, describing any steps to conceal the sequence until interventions were assigned | Methods – Randomisation and allocation concealment (immutable list in a password-protected platform; automated e-mail allocation) *(Manuscript: lines 257–259)* |
| **Implementation** | 19 | Whether the personnel who enrolled and those who assigned participants to the interventions had access to the random allocation sequence | Methods – Randomisation (study manager without prior participant contact; automated assignment, no access to the upcoming sequence) *(Manuscript: lines 258–259)* |
| **Blinding** | 20a | Who was blinded after assignment to | Methods – Randomisation (participants and staff not |

| | | | |
|---|---|---|---|
| | | interventions (e.g. participants, care providers, outcome assessors, data analysts) | blinded; data analysts were not blinded to group allocation; the primary endpoint is an objective laboratory biomarker) *(Manuscript: lines 260–262 (data analysts at line 262))* |
| | 20b | If blinded, how blinding was achieved and description of the similarity of interventions | Not applicable – open-label; no sham or placebo comparator |
| **Statistical methods** | 21a | Statistical methods used to compare groups for primary and secondary outcomes, including harms | Methods – Statistical analysis (robust ANCOVA; pre-specified gatekeeping strategy) *(Manuscript: lines 307–312)* |
| | 21b | Definition of who is included in each analysis (e.g. all randomised participants), and in which group | Methods – Statistical analysis (intention-to-treat by assigned group; n = 140 lipodia, n = 138 control) *(Manuscript: line 313; group sizes in Figure 1)* |
| | 21c | How missing data were handled in the analysis | Methods – Statistical analysis (multiple imputation under a missing-at-random assumption; jump-to-reference sensitivity analysis) *(Manuscript: lines 313–318)* |
| | 21d | Methods for any additional analyses (e.g. subgroup and sensitivity analyses), distinguishing prespecified from post hoc | Methods – Statistical analysis (pre-specified subgroups and sensitivity analyses; within-person responder analyses, with the 10% responder criterion post hoc and the LDL-C target criterion pre-specified) *(Manuscript: lines 321–328)* |
| **Results** | | | |
| **Participant flow (a flow diagram is strongly recommended)** | 22a | For each group, the numbers of participants who were randomly assigned, received intended intervention, and were analysed for the primary outcome | Results – Participant flow; Figure 1 (CONSORT flow diagram) *(Manuscript: lines 61–66; Figure 1 (lines 68–75))* |
| | 22b | For each group, losses and exclusions after randomisation, together with reasons | Results – Participant flow; Figure 1 (attrition at T1 and T2 per arm) *(Manuscript: Figure 1 (lines 68–75))* |
| **Recruitment** | 23a | Dates defining the periods of recruitment and follow-up for outcomes of benefits and harms | Methods – Trial design (first participant 26 Jan 2025; last T2 22 May 2026) *(Manuscript: lines 238–239)* |
| | 23b | If relevant, why the trial ended or was stopped | Trial ended as planned after the last participant |

| | | | |
|---|---|---|---|
| | | | completed T2 *(Manuscript: line 239)* |
| **Intervention and comparator delivery** | 24a | Intervention and comparator as they were actually administered (e.g. who delivered the intervention/comparator, how participants adhered, whether they were delivered as intended (fidelity)) | Results – Usage (uptake 99.3%; mean 38.4 min/week); self-guided delivery without clinician involvement *(Manuscript: lines 151–157 (usage); lines 263–276 (delivery))* |
| | 24b | Concomitant care received during the trial for each group | Methods – Control group and concomitant treatment (usual care in both arms; changes in lipid-lowering medication captured) *(Manuscript: lines 277–283)* |
| **Baseline data** | 25 | A table showing baseline demographic and clinical characteristics for each group | Results – Baseline characteristics; Table 1 *(Manuscript: lines 76–82 (Table 1 at line 82))* |
| **Numbers analysed, outcomes and estimation** | 26 | For each primary and secondary outcome, by group: the number included in the analysis; the number with available data at the outcome time point; the result for each group and the estimated effect size and its precision (e.g. 95% CI); for binary outcomes, both absolute and relative effect sizes | Results – Primary and Secondary endpoints; Tables 2–3 (adjusted differences, 95% CI, p, Cohen's d; responder proportions 41.7% vs 23.8%; target attainment 11.0% vs 6.2%) *(Manuscript: lines 89–108 (Table 2); lines 114–127 (Table 3))* |
| **Harms** | 27 | All harms or unintended events in each group | Results – Usage, satisfaction, and safety (adverse events; no adverse device effects) *(Manuscript: lines 157–159)* |
| **Ancillary analyses** | 28 | Any other analyses performed, including subgroup and sensitivity analyses, distinguishing pre-specified from post hoc | Results – Pre-specified subgroup analyses (age, sex, cardiovascular risk, baseline lipid-modifying medication; Supplementary Table S5 and Figure S1); jump-to-reference and per-protocol sensitivity analyses *(Manuscript: lines 133–149 (Suppl. Table S5/Figure S1 at line 149); sensitivity at lines 209 and 318)* |
| **Discussion** | | | |
| **Interpretation** | 29 | Interpretation consistent with results, balancing benefits and harms, and considering other relevant evidence | Discussion (mechanism; group-level MCID vs within-person response; benchmarking against other self-guided digital interventions) *(Manuscript: lines 160–212)* |

| | | | |
|---|---|---|---|
| **Limitations** | 30 | Trial limitations, addressing sources of potential bias, imprecision, generalisability, and, if relevant, multiplicity of analyses | Discussion (sample composition, local laboratories, open-label design, generalisability) *(Manuscript: lines 213–221)* |
